# ‘I don’t feel like I’m learning how to be a doctor’: early insights regarding the impact of Covid-19 on UK medical student professional identity

**DOI:** 10.1101/2021.08.01.21261101

**Authors:** Anna Harvey, Megan E L Brown, Matthew H V Byrne, Laith Alexander, Jonathan C M Wan, James Ashcroft, Nicholas Schindler, Cecilia Brassett

## Abstract

**Phenomenon:** Professional identity development is recognised as a core goal of medical education alongside knowledge and skill acquisition. Identity is a complex entity that can be conceptualised as externally influenced, but individually constructed. Integration from legitimate bystander to ‘old timer’ of the medical community of practice provides a backdrop for individual negotiation of professional identity. During Covid-19, the medical community of practice and education experienced significant disruption. We sought to investigate how these disruptions impacted professional identity development by examining conflicts between students’ identities highlighted by the pressures of the pandemic.

**Approach:** A mixed-methods survey was distributed to medical students in the UK. The survey was active from 2nd May to 15th June 2020, during the height of the first wave of the Covid-19 pandemic in the UK. Operating within the paradigm of constructivism, we conducted a reflexive thematic analysis of qualitative responses. Analysis was focused around the disruption to medical education, actions taken by medical students during this disruption, and the tension between student actions where they existed in conflict.

**Findings:** Three themes were constructed to describe the identities that participants felt were in conflict during the first wave of the Covid-19 pandemic: Status and role as a future doctor; status and role as a student; and status and role as a member of the wider community. Students noted that lack of clinical exposure was detrimental to their education, implicitly recognising that many aspects of professional identity formation are forged in the clinical environment. Participants were keen to volunteer clinically but struggled to balance this with academic work. Participants worried about the risk to their families and the wider community, and wanted to ensure that their skills would add value to the clinical environment. Volunteers felt frustrated when they were unable to perform tasks which aligned with their identities as future doctors, with the exception of participants who worked as interim FY1s (FiY1s), which aligned well with the roles of FY1s.

**Insights:** As hypothesised, the participants in this study experienced disruptions to their professional identity development during the first wave of the Covid-19 pandemic in 2020. This work provides early evidence, collected at the beginning of the pandemic, that the effects of disruptions to professional identity development were wide-reaching, often negative, and represent an important topic for future exploration. Given that the pandemic has highlighted areas of identity tension, these findings have the potential to provide insight into how medical training can better nurture professional identity development during and beyond international crises.

## Introduction

Professional identity development in medicine describes the process by which students move from being a “member of the public” to a member of the profession. Holden et al. described professional identity formation as the “transformative journey through which one integrates the knowledge, skills, values, and behaviours of a competent, humanistic physician with one’s own unique identity and core values”^1^. Developing a strong professional identity is considered a key facet of medical education. Indeed, evidence suggests that those with strong professional identities achieve higher levels of well-being and success in their careers as doctors ^2^. Medical students and doctors have multiple identities, and it is the successful integration of existing identities with the new identity of “professional” that represents the goal of professional identity development in medical education ^3^.

Professional identity is developed in part through interactions with the formal, informal, and hidden curricula ^3^. The formal curriculum represents the overt teaching and learning that medical students experience, as in anatomy, physiology, and clinical teaching. The informal curriculum represents learning undertaken outside of the structured class or placement environment. The hidden curriculum consists of the overarching structural and cultural “rules” of medicine - what might be thought of as medicine’s “etiquette” ^4–6^. Creuss et al. summarise the factors that contribute to this process of professional identity development, detailing a functionalist approach to the theory of socialisation, whereby students become part of the medical profession through role modelling, conscious and unconscious reflection on experiences, as well as participation in cultural rituals and engagement with symbols ^7,8^. Professional identity development is a complex process that can be conceptualised from a psychological orientation as individually mediated. Individuals construct professional identities that align with their own personal values, beliefs and experiences, which in turn are influenced by external factors ^7^. Learning through these three curricula results in students becoming increasingly integrated into the medical community of practice, membership of which provides the external backdrop for individual identity development. Communities of practice theory as applied to medicine conceptualises the profession as a group that is “engaged in a common activity” - the practice of medicine - where members are united in shared activities, overlapping knowledge, and a set of shared values, beliefs and heritage focused on a mutual enterprise ^9^. Communities of practice theory provides a framework within which individuals develop their professional identities as they move from the periphery of a community towards full membership and ‘old-timer’ status, learning how to “think, act and feel like a physician” as they progress ^10^.

During the Covid-19 pandemic, all three curricula (formal, informal and hidden) were disrupted by reformulations to university studies and clinical placements. The formal curriculum shifted online, with relative success in transforming didactic teaching and assessment into digital formats ^11–13^. By moving these sessions online, and, in many cases, removing students from the clinical environment ^12^, medical students had less access to informal conversations with their peers and seniors. The structures, social norms and culture that comprise the hidden curriculum to which medical students are usually exposed were also disrupted, if not irrevocably changed, by such an international crisis. This interruption and adaptation to medical education would have subsequently interrupted opportunities for medical students to integrate into the medical community of practice ^3^.

There is a significant body of previous work exploring how and why professional identity develops during the course of medical school, but as yet fewer studies have investigated the impact of Covid-19 on medical students’ professional identity development. It has been postulated that the changes to the learning environment caused by Covid-19 may have changed the process of professional identity development and integration into the medical community of practice ^14^, though little qualitative data exists to support this claim, a gap this study intends to address. Some early work from clinical educationalists has suggested alternative routes to support professional identity development during Covid-19, calling for a “reimagining” of professional identity through, for example, work as an allied health professional, public health or community work, and through social media use ^15^. One US institution even used the pause in clinical activity to deliver a structured professional identity curriculum to their medical students ^16^. One study of Indonesian students’ reflections on their role during Covid-19 focused on how students have adapted their learning to the new demands of the pandemic and argued that, despite disruptions, students recalled experiences that had enabled continuation of their professional identity development ^17^. This study is not intended to replace the calls for a reimagining of how student doctors integrate their identities with those of a professional, but rather to explore how disruptions influenced identity development, particularly focusing on the conflicts between the different identities that medical students possess. As has been explored in literature regarding dissonance between professional and personal identities, such as Costello’s work regarding race, class, gender and professional identity, conflict between different facets of identity can affect the wellbeing and career success of those in the process of integrating into a profession, which is why we have focused on conflict here ^18^.

In this study, we explore early indicators of how the process of professional identity development was disrupted or changed due to Covid-19 and the subsequent changes made to the medical curriculum. We hypothesised that, with the removal of students from clinical and academic environments during Covid-19, opportunities to experience the facets of the formal, informal and hidden curricula that facilitate professional identity development may have been disrupted or changed, thus affecting an individual’s professional identity development. As disruptions due to Covid-19 may continue in the form of blended or even fully online delivery of medical education, an investigation of the challenges posed by recent disruptions on professional identity development has benefits beyond the current pandemic.

## Methods

The STROBE guidelines for cross-sectional studies were followed ^19^. The study was an online survey that was distributed to medical students studying at all UK medical schools. Ethical approval for the study was obtained from the University of Cambridge Psychology Research Ethics Committee (Approval number: PRE.2020.040).

We distributed a mixed methods survey to UK medical students. Quantitative data from the survey will be published elsewhere. Inclusion criteria were any medical student studying at a UK medical school, including those who may have graduated early this year due to Covid-19. Medical schools were identified by their listing on the UK Medical Schools Council website ^20^. All medical schools were contacted via their medical school office general enquiries email and the Dean responsible for education and asked to distribute the survey. Messages were also posted once weekly to social media (via Twitter and Facebook accounts set up specifically for the purposes of promoting the study and to medical student university groups) asking medical students to complete the questionnaire and share the survey to recruit participants using a snowball approach.

The survey consisted of 53 questions assessing (quantitatively and qualitatively) previous clinical experience, attitudes to volunteering and motivation and barriers, volunteering role, medical education, issues currently faced, and safety. Survey development was informed by a systematic review of existing literature on volunteering during pandemics and disasters^21^. Questions were then developed from previously used scales by MHVB and JA with expert input and consultation with medical students, and final questions were reviewed by medical students to establish face validity.

The responses to three qualitative questions (“If you have ethical concerns what are they?”, “What issues are you currently facing as a medical student during the coronavirus pandemic?”, “Have you heard any stories about medical students volunteering experiences? Please describe them.”) were used for this work. The survey is available in the supplementary material.

### Context

The survey was active from 2nd May to 15th June 2020, providing a snapshot of the experiences of a wide variety of students in the midst of the disruption. The survey was distributed during the height of the first wave of the Covid-19 pandemic in the UK. The Medical Schools Council issued guidance on the 13th March 2020 indicating that training of medical students should be based on local needs, and that final year students who had met the GMC Outcomes for Graduates would be given provisional registration earlier than usual ^22^. Practically, this meant that most students were removed from the clinical environment in their capacity as medical students, with some having the opportunity to volunteer in roles both within and outside the clinical environment. These roles were allocated locally and there was considerable diversity in the roles students were able to take up. For graduands, a structured “Interim FY1 programme” was introduced, aligned with the job role of a Foundation Year 1 doctor, including provisional registration with the GMC ^23^.

### Research approach

Operating within the paradigm of constructivism, we conducted a reflexive thematic analysis following Braun and Clarke’s six-step approach to code and construct themes ^24^. In this context, identity was conceptualised as an individually mediated but dynamic entity influenced by social relationships, and constructed, in part, through role and through increasing levels of integration into the medical community of practice, as discussed and defined in the introduction. Given the range of viewpoints and backgrounds of respondents represented in the existing data set, this methodological approach is appropriate in that it allows for different interpretations and constructions of identity to sit alongside one another, allowing for examination of tensions between the multiple facets of one’s identity. Analysis was focused around the disruption to medical education, actions taken by medical students during this disruption, and the tension between student actions where they existed in conflict.

Three authors (LA, MHVB and AH) familiarised themselves with the data by reading and re-reading the data. Initial ideas were noted. LA and MHVB created initial descriptive codes using an inductive approach, and the data set was coded systematically. To identify themes, a semantic approach was used. Codes were then analysed for patterns, grouped, summarised, interpreted and discussed with all authors to generate early themes. The codes and their relation to the data and themes were then systematically reviewed by AH in light of the conceptual framework discussed previously, and with a particular focus on identity formation and conflict. Three themes were constructed. Themes and subthemes were checked against the data set as a whole and refined. Themes and subthemes were discussed at length by all authors, and a final set of themes named and defined. To preserve participant voice, quotes were selected to illustrate the final themes and highlight areas of concordance and tension between them.

### Reflexivity statement

Reflective notes were kept to promote interrogation of the relationship between authors and the dataset. Given our constructivist approach, we acknowledge that realities are variable, and that each researcher brings their own reality to the dataset, influencing interpretation. Rather than viewing this as an issue, the authors perceive this as a strength - their insider statuses, background in identity theory, and education offer a unique perspective that has deepened analysis.

AH is a medical student who spent time during the Covid-19 pandemic reporting on issues faced by other students. MELB is a PhD student in medical education, previously familiar with identity development theory. MHVB and JA are Academic Clinical Fellows who worked in a clinical capacity with volunteer medical students during the Covid-19 pandemic and have postgraduate qualifications in medical education, LA and JCMW were final year medical students during the Covid-19 pandemic, NS is an NHS consultant and tutor in medical education with a research interest in postgraduate medical training. CB holds multiple senior medical educational university positions and has been directly involved in the changes being made to medical student teaching during the COVID-19 pandemic.

### Team reflexivity

This study was conducted by a diverse collaborative of medical students, doctors in training, medical education researchers, and consultants spanning multiple educational institutions. This plethora of viewpoints and independence from any single institution allowed the authors a broad spectrum of thought in relation to the data and mitigates the temptation to overemphasise or dismiss contributions made by participants.

## Results

### Respondent demographics

1145 medical students completed the survey. Of these, 928 provided qualitative responses analysed in this study. 687 out of 928 (74.0%) respondents were female. The median age of respondents was 22 (interquartile range, IQR, 20-24), as shown in Figure 1. The modal year group of the respondents was third year, and 129 (14.0%) were in their final year of study. At the time of responding (median response date 16/04/2020), 282 out of 928 (30%) had commenced volunteering in a clinical capacity.

**Figure 1:**
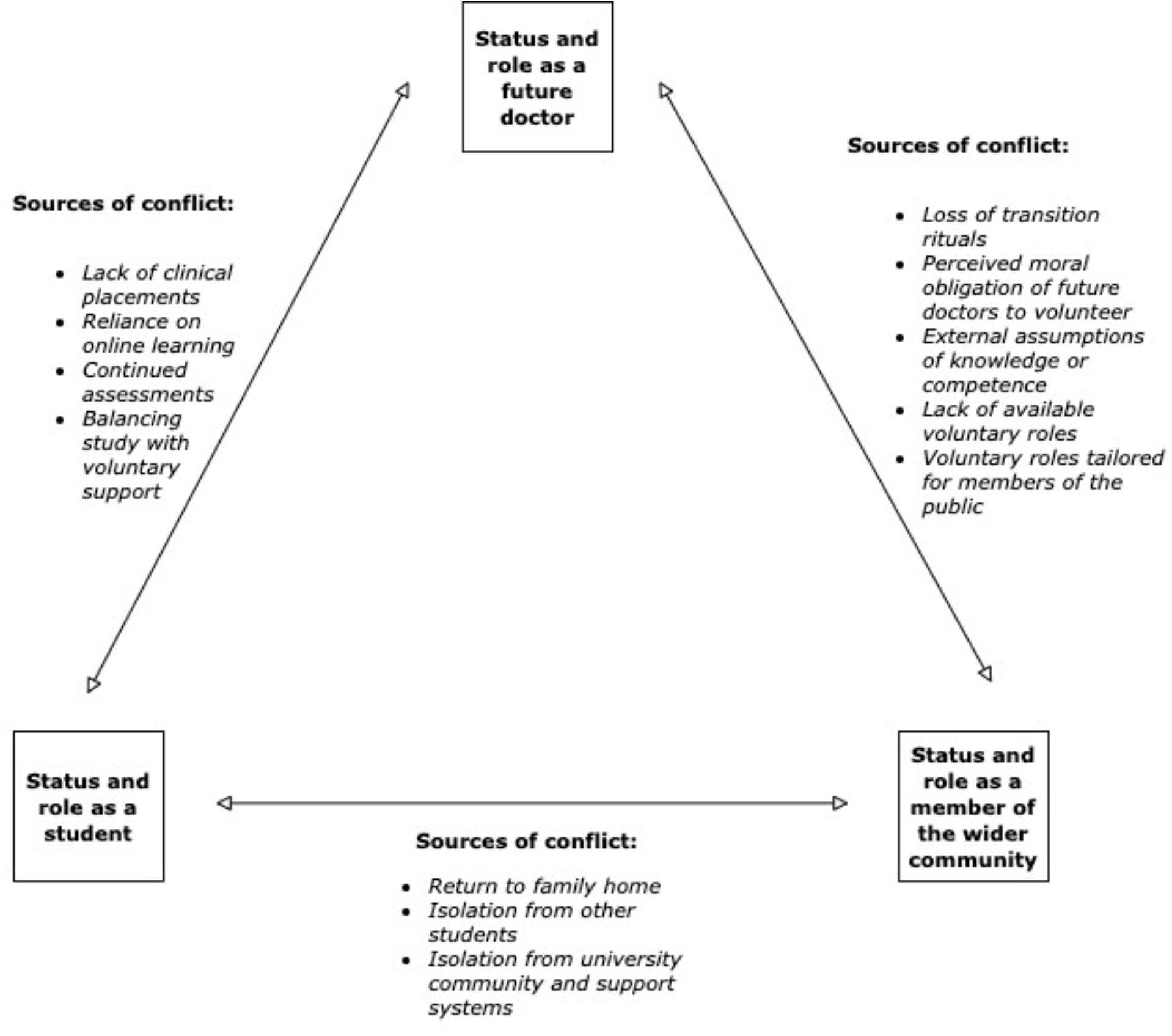
Identity themes and the conflicts between them as expressed by our participants.

### Thematic analysis

Three themes were constructed to describe the identities that participants felt were in conflict during the first wave of the Covid-19 pandemic:

- Status and role as a future doctor
- Status and role as a student
- Status and role as a member of the wider community

Figure 1 provides an overview of the themes and conflicts between them.

Participants from all years of all medical schools identified a lack of opportunity to learn within a clinical environment as a key educational issue, highlighting that “nothing can really replace face to face patient contact” (Participant 163). Students implicitly identified that “becoming a doctor” involves more than simply learning material didactically: “no clinical skills, no hospital or lab experience…I don’t feel like I’m learning to be a doctor” (Participant 608). Participants worried about the cancellation of assessments in regard to academic competency when moving into their next year of study, sometimes linked to the lack of clinical exposure: “missing placement blocks and learning, not doing exams and therefore not knowing if I’m competent” (Participant 348). Transition symbols such as end of year assessments were seen as important by participants in assuring their competence to move into the next stage of their medical school careers, and continuing their integration into the community of practice.

For final year medical students, many of whom were moved into the workforce early, the loss of graduation rituals left participants dissatisfied with the legitimacy of their transition into qualified professionals, with “no elective, no free time, no graduation ceremony, no final year dinner, no closure from the hardest years of my life at medical school” (Participant 42), with some stating that the lack of these rituals left them with “impostor syndrome” (Participant 19). Others felt that being unable to sit their final examinations, or undertaking truncated versions, caused them to doubt their skills as a junior doctor: “my medical degree ended so abruptly and I didn’t get to take my finals which means I have not revised as intensely as I would have otherwise, therefore may lack knowledge/skills” (Participant 180).

Participants were keen to volunteer in a clinical capacity as their placements and often assessments had been cancelled, though many found it challenging logistically to secure roles. Many who did secure roles as support workers or similar outlined the “great benefit in many ways - learnt new communication skills I otherwise would not have developed at medical school, part of team, clinical experience” (Participant 375). Many participants valued the opportunities to work in allied roles, and appreciated the additional insight this would bring to their future medical careers: “working as HCSW [healthcare support worker]…I have been able to understand the patient experience a lot better and understand how the different roles of a HCSW, nurse and doctor all fit together with regards to patient care. My inter-professional skills have also improved. As a medical student it can be difficult to understand how you fit in this dynamic but being able to work as HCSW has enlightened me in the reality of the daily ins and outs” (Participant 1024).

In contrast, others did secure roles, but felt that their skill sets were not adequately used, mentioning that they were unable to undertake some skills they felt comfortable with: “for example, I am currently not allowed to cannulate or do bloods even though that would be the most helpful thing…so it doesn’t appear the roles have been thought through” (Participant 757). Many linked their roles with their “utility” - as a medical professional in training, they became frustrated when their roles were not aligned with skills in which they were competent. Those skills often aligned more closely with the role of a junior doctor than, for example, a healthcare assistant. Some participants drew comparisons with allied health students, who were more often kept in the clinical environment, feeling “that medical students are not useful or recognised to the NHS…all my allied health professional friends have recognised roles. We do not, we are very much not wanted I feel” (Participant 232), with others highlighting that pandemic pressures had highlighted existing tensions with medical students being unable to properly integrate into clinical teams, with “the pandemic [proving] that medical students are the bottom rung of the ladder and their placements aren’t important, as soon as anything happens we are seen as a nuisance” (Participant 318). Concerningly, some participants reported a degree of perceived hostility from other members of the healthcare team. There was additional guilt around this, especially in relation to being paid. These experiences reflect medical students’ perceptions that other healthcare professionals do not regard them as “members” of the wider healthcare team, and that medical students are unable to add value or be useful: “[allied health professionals] complained to us that we were doing work that they could do and were more qualified to do, while they had very little to do, and thus they felt that paying us was wasting NHS funding” (Participant 514).

Perhaps unsurprisingly, guilt was expressed by many participants, particularly where conflicts existed between the actions demanded by different aspects of their identities. For example, there was significant guilt described around voluntary work - participants felt that as future doctors they had an obligation to utilise their skills, but were unsure of their relative usefulness balanced with the risk to themselves and their wider communities: “I felt like I had a duty to help, as in 3 years’ time I would be working as a doctor anyway. I have skills and understanding that other members of the public don’t have and I should use that for the greater good. I wanted to help so much that I did apply to work in a hospital until my family circumstances changed” (Participant 1114). Another key concern was “weighing up study and revision and helping in the ICUs” (Participant 242), especially with “not clear enough guidance from the medical school; dilemma for volunteering vs studying for exams” (Participant 959).

Many of these dilemmas were compounded by students returning to their family homes during the first lockdown period, and “trying to continue with medical school studies at home,” (Participant 100) some with additional pressures of household work or vulnerable family members. Isolation from medical school friends and support systems left some participants “trying to maintain academic motivation without the support of colleagues” (Participant 470), and others simply “missing friends and normal life” (Participant 79). Interestingly, some participants reported that family members had high expectations of their medical student relatives, mentioning “pressure from family to volunteer as they believe I am more qualified than I am” (Participant 228). Others reported feeling pressured to explain rapidly-changing public health measures, “people expecting me to know more than I do about the pandemic and having to answer friends’ and colleagues’ questions continually about this; this is incredibly stressful” (Participant 370). Guilt and frustration were expressed where these external perceptions of a student’s identity and membership of the profession conflicted with a student’s own perceptions of their position and ability as a healthcare professional in training.

A key exception to the issues with voluntary work outlined above was the group of participants who worked as interim FY1s, aligned with the role of an FY1. Whilst these newly-graduated doctors had missed out on the traditional transition rituals, the extended “shadowing” period was effective in supporting their new identities as junior doctors, as well as relieving the usual pressures of the first weeks and months of working: “frankly, I feel it’s made been hugely beneficial as the work is pretty easy and manageable, and has given me time to get to grips with what being a newly registered doctor is like with 0% of the usual responsibilities yet 100% of the pay” (Participant 436).

**Table 1:**
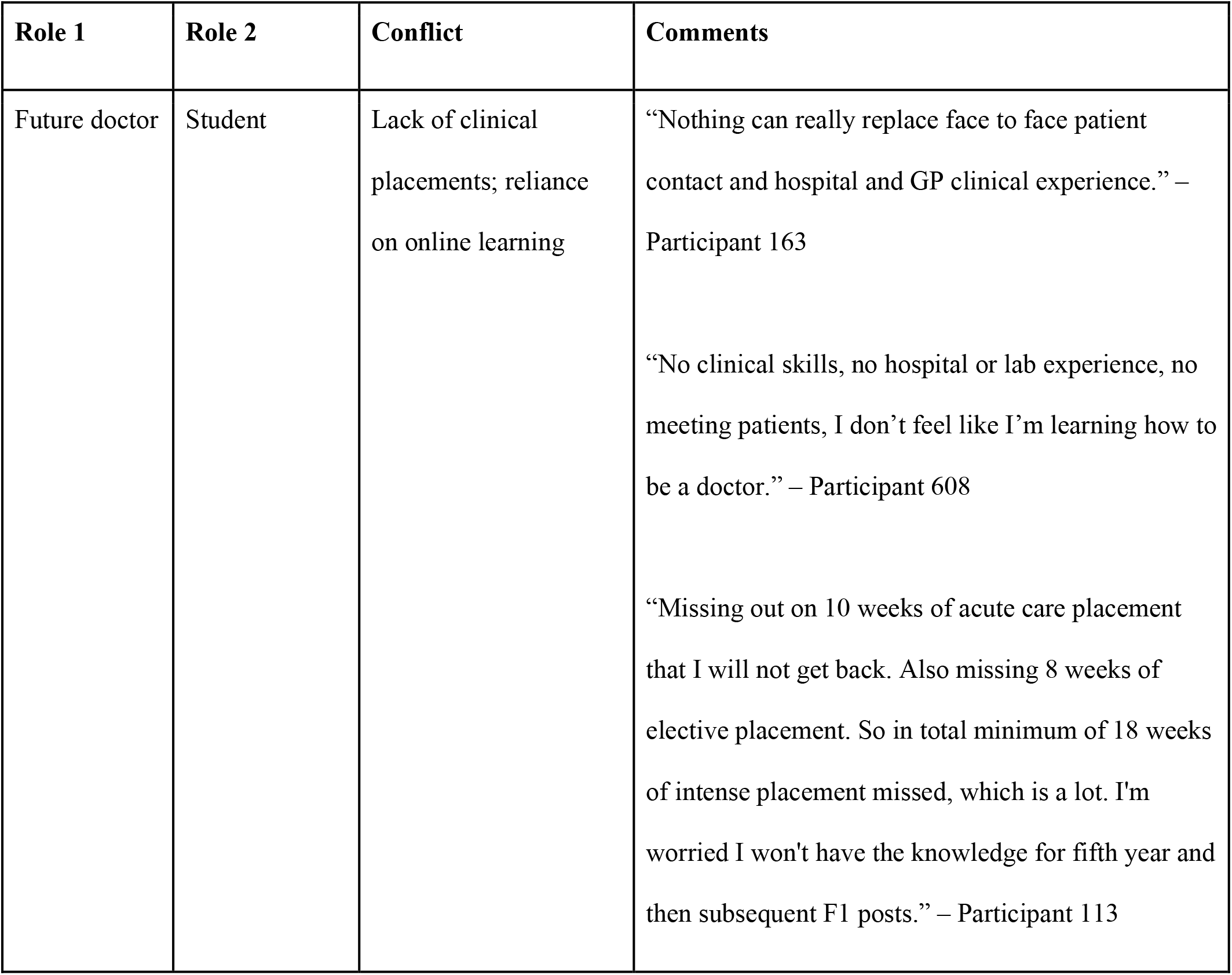
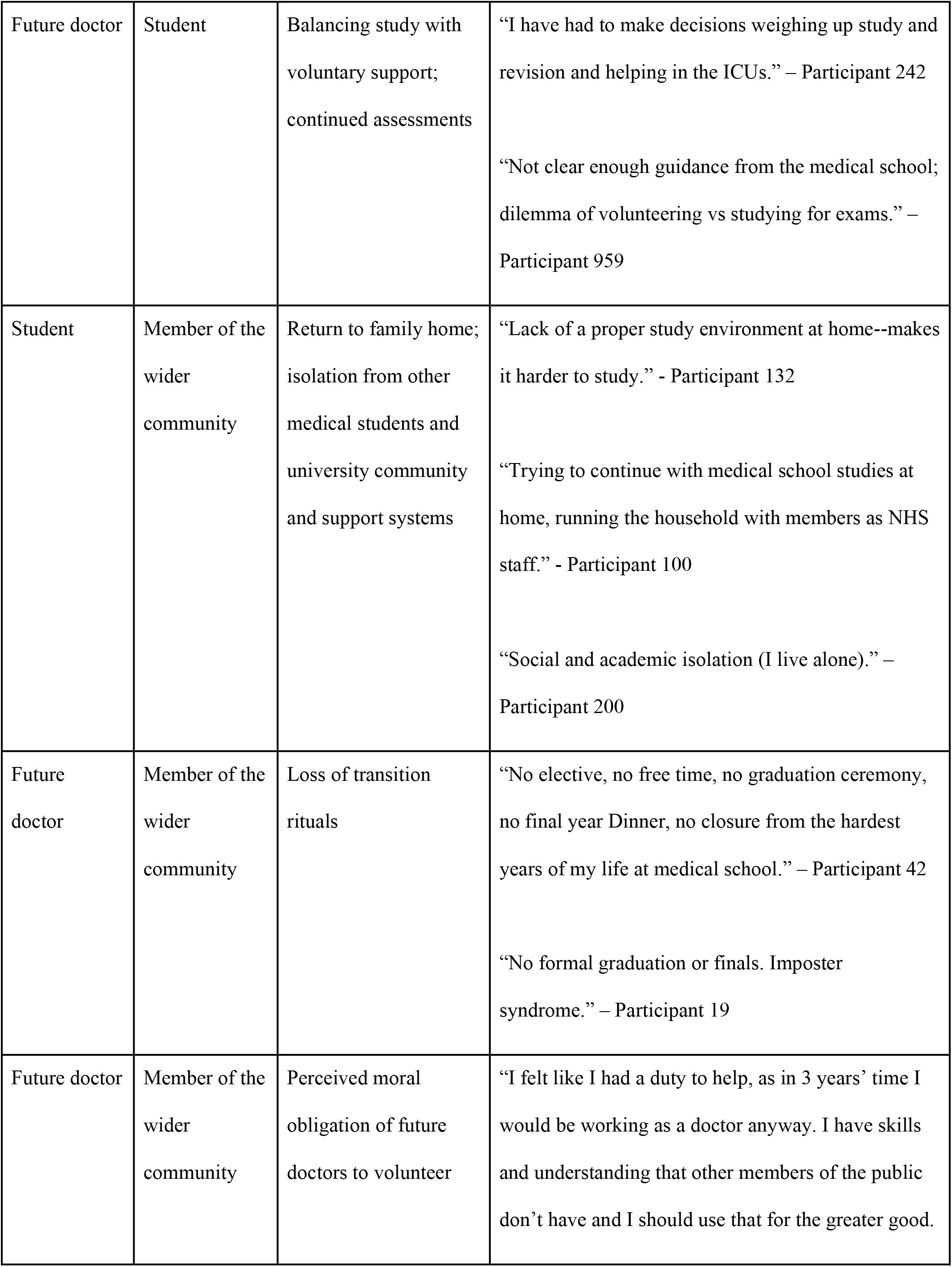
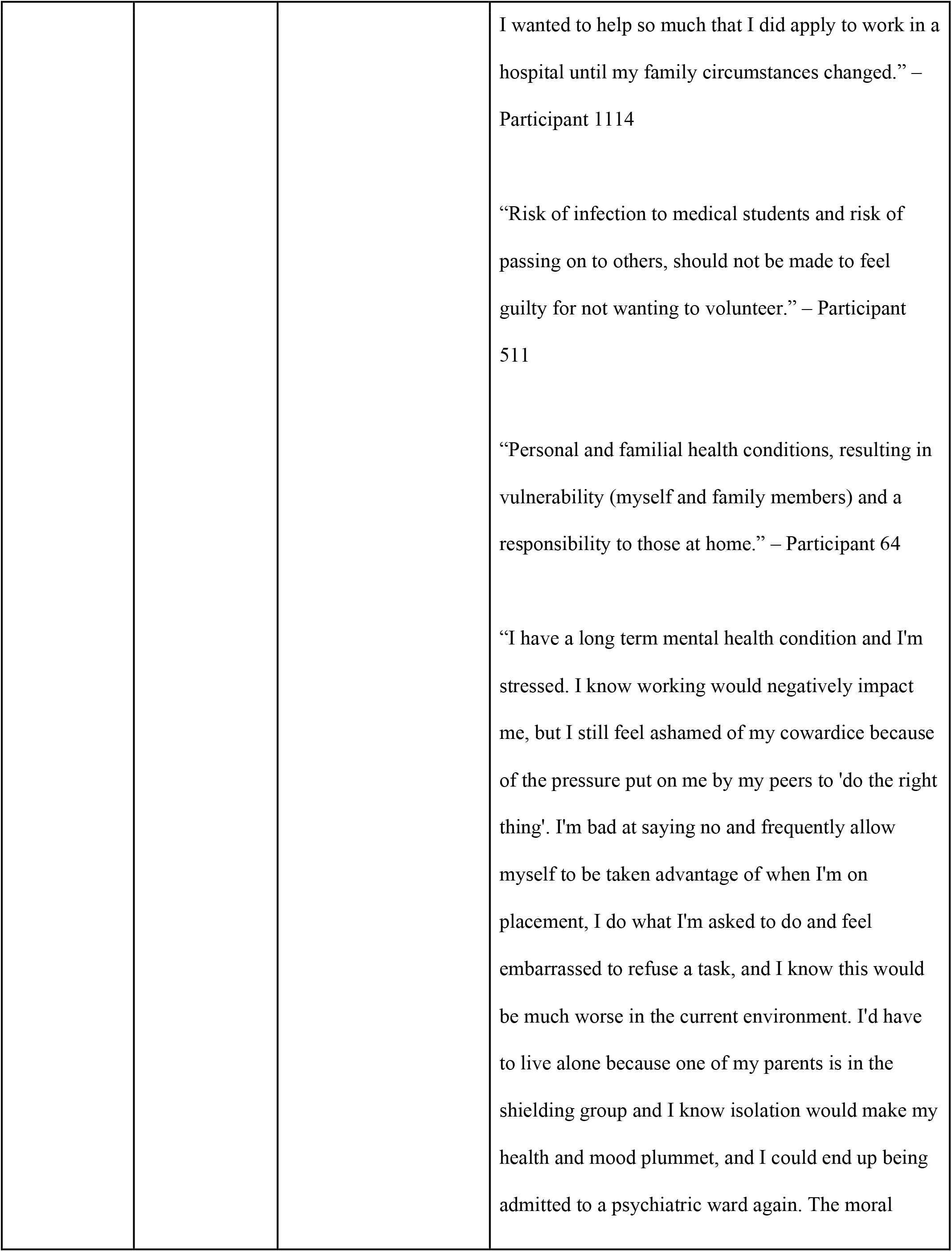
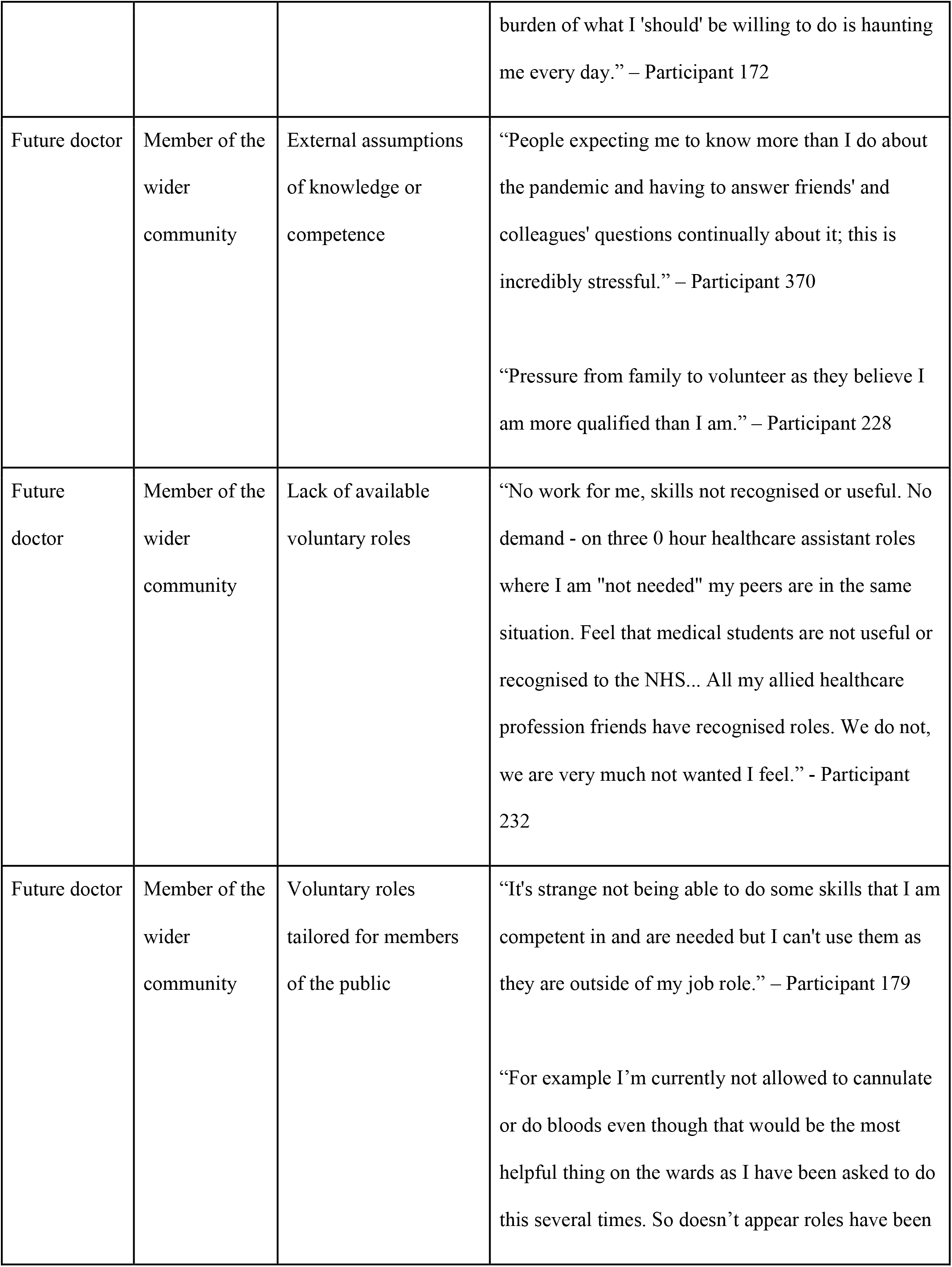
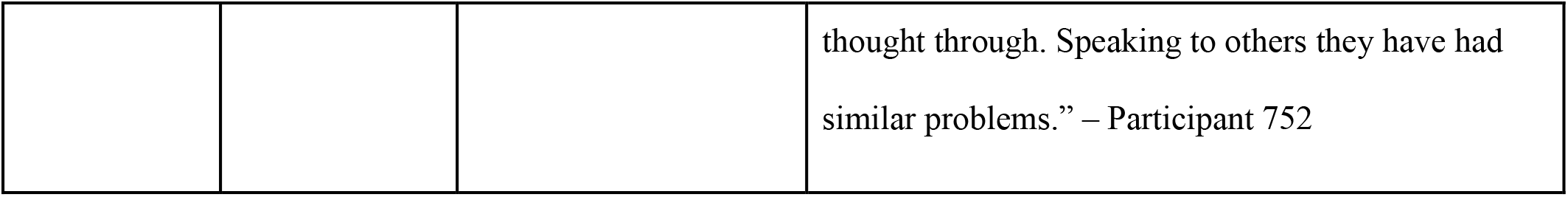
Summary of the conflicts between the identity themes constructed from the data with representative quotes from participants

## Discussion

As hypothesised, the participants in this study experienced disruptions to the usual process of professional identity development during the first wave of the Covid-19 pandemic in 2020. This work provides early evidence, collected at the beginning of the pandemic, that the effects of disruptions to professional identity development is an important topic for future exploration. This has the potential to provide insight into how medical training can better nurture professional identity development, as the pandemic has highlighted areas of tension.

Some work has suggested that medical students’ professional identity formation is driven in part by both “professional inclusivity” – doctors and other healthcare professionals treating students as future doctors – and “social exclusivity” – medical students considering themselves as socially distinct from other groups of students ^25^. During Covid-19, casual and informal interactions between groups of medical students were reduced, and many students returned to their family homes. As a consequence, both professional inclusivity and social exclusivity were reduced, and this potentially weakened students’ professional identity development. The key exception to this was the group of students who graduated early to take up interim FY1 posts. In this cohort of respondents, who had roles aligned to those of a junior doctor, feedback was positive, which is supported by some early insights into the interim Foundation posts in the literature ^26^. As well as structured roles, interim FY1 doctors were immersed in the milieu of their jobs, including social contact with other junior doctors, which may also contribute to strengthening their professional identity, in contrast to the challenges to professional identity development experienced by more students from junior years. For new graduands, it was the loss of transition events and rituals, such as final exams, that was cited as a source of anxiety, with participants concerned that the loss of assessments meant they would be poorly prepared for their jobs as junior doctors.

Medical students self-categorise as future doctors and, even at the very beginning of their medical degrees, are likely to describe themselves in line with attributes associated with doctors, who are a large and well-known group ^27^. During Covid-19, medical students have been, in some ways, dislocated from their usual anchors to the profession and have developed new behaviours in which to anchor their perceptions of what a future doctor “should’’ be doing in a crisis, often influenced by their perceptions of external influencers such as parents and friends. For many, this meant volunteering in a clinical capacity. Our data highlighted the conflicts between students’ identities, and the difficulties some participants had in resolving these conflicts. As noted by Frost et al., tension between different aspects of identity exists within the usual course of medical studies. They describe students resolving these tensions in different ways: some by aligning more strongly with standardisation, and others by rejecting standardisation and focusing on how their identities outside of medicine mark them as different from others in the community ^28^. Under the specific circumstances of our respondents, resolving these tensions in the way described by Frost was potentially more challenging, as the uncertainties of the pandemic meant there was no “standard” with which to align themselves.

Whilst the hidden curriculum and the lack of opportunities to learn “implicit” skills were not explicitly considered by respondents (indeed, by its very nature, the hidden curriculum is just that-hidden), there is recognition within our data that one does not “become a doctor” through lectures on an online platform alone. Some participants’ comments implied that for them the risk/benefit analysis was in favour of seeing patients rather than potentially lacking the skills to practise effectively as a doctor when they graduate. Preparedness for practice is still a key priority in UK medical education, and there is extensive literature suggesting that newly qualified foundation doctors lack preparedness even after completing their medical education in ‘the usual way’ ^29–31^. It is reasonable that medical students would be concerned about the impact of this disrupted education on their future competency as junior doctors. This mismatch of expectation, and lack of guidance around what to prioritise, left respondents questioning their own alignment with the profession and status as a future doctor, especially if they were unable to volunteer clinically.

Those who were able to find voluntary posts valued volunteering as helping them to develop personally and professionally. Many participants enjoyed and valued their experiences of working in different types of roles, being more involved in direct patient care and feeling part of a team. Conversely, some respondents felt their skills as a medical student had been under-utilised. This mismatch between the roles given to medical students, and their perception of their identity as a future doctor (often conceptualised as a leader or decision maker), left some respondents feeling that their professional identity had been undermined – they were not being ‘professionally included’ and regarded as future doctors by other members of the healthcare team. Role recognition is considered an important facet of developing a robust professional identity ^32^. Indeed, for other professions – such as Physician Associates – lack of role recognition amongst other healthcare professionals has been noted to hinder the development of professional identity ^33^. Some respondents compared their positions to those of student nurses and other allied health professionals, who were given the option to remain in the clinical environment in their usual placement structure. As there has been very little work on the perceptions and experiences of such allied health students during Covid-19, it is interesting to consider how experiences might differ. One study of qualified nurses in China found that the Covid-19 pandemic solidified nurses’ professional identity, with a sense of professional unity and utility to the public noted to be particularly influential ^34^. This early work stands in contrast to our findings regarding the experiences of many medical students, who were often removed from their peer groups, struggling to find roles in which they felt “useful.”

Our results highlight early signals that the disruption caused by Covid-19 had an impact on the professional identity development of our participants. Further research is needed to explore this in greater depth. Longitudinal work could explore how this may change students’ perceptions of their own identities and position within the medical profession, especially the differences between those who did and did not take on clinical voluntary work. Previous work on strategies to “effectively scaffold the necessary critical reflective learning and practice skill set for our learners to support the shaping of a professional identity” has highlighted the importance of structured reflective practice in “teaching” professional identity to medical students ^35^. Small group work where students are invited to share personal narratives which are then discussed and fed back by trained education professionals, including members of the multidisciplinary team, have reported success in student feedback.

Universities that do not already facilitate such groups should consider integrating such strategies into their curricula to help students continue their identity development as curricula are adapted to Covid-19 restrictions, and beyond.

## Limitations

The nature of the collection and analysis of the qualitative data used in this paper may limit the transferability of the conclusions. For example, just under two thirds of our respondents identified as female, though women comprise 55% of the medical student population ^36^.

As the qualitative data were collected via a short form survey, we were unable to clarify or seek further depth in the free text responses that have been thematically analysed in this paper. Responses were self-reported with little guidance for the structure and depth of answers, resulting in variable levels of engagement with reflection upon respondents’ experiences ^37^. We selected this method over in-depth interviewing for pragmatic reasons, as a survey-based data collection method was better suited to reflect the varied experiences of medical students during the crisis, rather than retrospectively.

## Conclusion

The spring 2020 wave of the Covid-19 pandemic and its disruption to medical education in the UK has implications for the professional identity development of medical students. The significant break in studies, lack of transition rituals, and the perceived under-utilisation of medical students’ skill sets have all served to disrupt students’ identity development. Notably, we found that those students who were allocated a role aligning with their pre-conceived professional identity, *i*.*e*. – interim FY1 roles, reported far less conflict and guilt than more junior students, many of whom were unable to secure roles which they felt aligned with their skill sets and identity. Students recognise that learning to be a doctor requires more than online learning, and that experience in the clinical environment is not only desirable but is essential to development of their professional identity. Universities should consider how their online programmes of learning, which will continue into the 2020-21 academic year, may be affecting the process of identity development. This particularly needs to be considered when planning learning for students early in their medical school careers, for whom clinical experience might be regarded as less important and potentially dispensable. Further research is needed to continue to explore how students negotiated the challenges to their professional identity development during Covid-19.

## Supporting information

Supplemental Survey Questions

## Data Availability

None

## References

1. Holden MD, Buck E, Luk J, et al. Professional identity formation: creating a longitudinal framework through TIME (Transformation in Medical Education). Acad Med. 2015;90(6):761–767. doi:10.1097/ACM.0000000000000719

2. Monrouxe L V. Identity, identification and medical education: why should we care? Med Educ. 2010;44(1):40–49. doi:10.1111/j.1365-2923.2009.03440.x

3. Goldie J. The formation of professional identity in medical students: considerations for educators. Med Teach. 2012;34(9):e641–8. doi:10.3109/0142159X.2012.687476

4. Hafferty FW, Franks R. The hidden curriculum, ethics teaching, and the structure of medical education. Acad Med. 1994;69(11):861–871. doi:10.1097/00001888-199411000-00001

5. Lempp H, Seale C. The hidden curriculum in undergraduate medical education: qualitative study of medical students’ perceptions of teaching. BMJ Br Med J. 2004;329(7469):770. doi:10.1136/BMJ.329.7469.770

6. Hundert EM, Hafferty F, Christakis D. Characteristics of the informal curriculum and trainees’ ethical choices. Acad Med. 1996;71(6):624–642. doi:10.1097/00001888-199606000-00014

7. Cruess RL, Cruess SR, Boudreau JD, Snell L, Steinert Y. A schematic representation of the professional identity formation and socialization of medical students and residents: a guide for medical educators. Acad Med. 2015;90(6):718–725. doi:10.1097/ACM.0000000000000700

8. Brown MEL, Finn GM. When I say… socialisation. Med Educ. February2021:medu.14469. doi:10.1111/medu.14469

9. Barab SA, Barnett M, Squire K. Developing an Empirical Account of a Community of Practice: Characterizing the Essential Tensions. J Learn Sci. 2002;11(4):489–542. doi:10.1207/S15327809JLS1104_3

10. Pollak O. Robert K. Merton, George G. Reader and PATRICIA L. Kendall (Eds.). The Student-Physician: Introductory Studies in the Sociology of Medical Education. (A report from the Bureau of Applied Social Research, Columbia University.) Pp. xii, 360. Cambridge, Mass. Ann Am Acad Pol Soc Sci. 1960;328(1):207–207. doi:10.1177/000271626032800167

11. Torres A, Domańska-Glonek E, Dzikowski W, Korulczyk J, Torres K. Transition to online is possible: Solution for simulation-based teaching during the COVID-19 pandemic. Med Educ. 2020;54(9):858–859. doi:10.1111/medu.14245

12. Arora A, Solomou G, Bandyopadhyay S, et al. Adjusting to Disrupted Assessments, Placements and Teaching (ADAPT): a snapshot of the early response by UK medical schools to COVID-19. medRxiv. 2020:2020.07.29.20163907. doi:10.1101/2020.07.29.20163907

13. Kanneganti A, Sia C-H, Ashokka B, Ooi SBS. Continuing medical education during a pandemic: an academic institution’s experience. Postgrad Med J. May 2020. doi:10.1136/postgradmedj-2020-137840

14. Kinnear B, Zhou C, Kinnear B, Carraccio C, Schumacher DJ. Professional Identity Formation During the COVID-19 Pandemic. J Hosp Med. 2021;16(1):44–46. doi:10.12788/jhm.3540

15. Stetson G V., Dhaliwal G. Using a time out: Reimagining professional identity formation after the pandemic. Med Educ. November 2020:medu.14386. doi:10.1111/medu.14386

16. Stetson G V., Kryzhanovskaya I V., Lomen-Hoerth C, Hauer KE. Professional identity formation in disorienting times. Med Educ. 2020;54(8):765–766. doi:10.1111/medu.14202

17. Findyartini A, Anggraeni D, Husin JM, Greviana N. Exploring medical students’ professional identity formation through written reflections during the COVID-19 pandemic. J Public health Res. 2020;9(Suppl 1):1918. doi:10.4081/jphr.2020.1918

18. Costello CY. Professional Identity Crisis: Race, Class, Gender, and Success at … - Carrie Yang Costello - Google Books. https://books.google.co.uk/books?hl=en&lr=&id=YDpnjmISv1kC&oi=fnd&pg=PP9&dq=costello+identity+dissonance&ots=wH55DulwZJ&sig=ih-gpxDlgSrDo2QleYhsT4778F8#v=onepage&q=costelloidentitydissonance&f=false. Accessed June 29, 2021.

19. STROBE Initiative. STROBE Checklist for cross-sectional studies.

20. Medical schools v Medical Schools Council.

21. Ashcroft J, Byrne MH V, Brennan PA, Davies RJ. Preparing medical students for a pandemic: a systematic review of student disaster training programmes. Postgr Med J. 2020;0:1–12. doi:10.1136/postgradmedj-2020-137906

22. Atherton J. Advice from Medical Schools Council to UK Medical Schools on Actions Surrounding Covid-19.

23. Joint statement from the UK Health Departments, the General Medical Council, Health Education England, NHS Education for Scotland, Health Education and Improvement Wales, the Northern Ireland Medical and Dental Training Agency and the MSC. Joint Statement from the UK Health Departments, the General Medical Council, Health Education England, NHS Education for Scotland, Health Education and Improvement Wales, the Northern Ireland Medical and Dental Training Agency, and the Medical Schools Cou.; 2020.

24. Braun V, Clarke V. Using thematic analysis in psychology. Qual Res Psychol. 2006;3(2):77–101. doi:10.1191/1478088706qp063oa

25. Weaver R, Peters K, Koch J, Wilson I. “Part of the team”: professional identity and social exclusivity in medical students. Med Educ. 2011;45(12):1220–1229. doi:10.1111/j.1365-2923.2011.04046.x

26. Youssef S, Zaidi S, Shrestha S, Varghese C, Rajagopalan S. First impressions of the foundation interim year 1 postings: positives, pitfalls, and perils. Med Educ Online. 2020;25(1):1785116. doi:10.1080/10872981.2020.1785116

27. Burford B, Rosenthal-Stott HES. First and second year medical students identify and self-stereotype more as doctors than as students: a questionnaire study. BMC Med Educ. 2017;17(1):209. doi:10.1186/s12909-017-1049-2

28. Frost HD, Regehr G. “I AM a Doctor.” Acad Med. 2013;88(10):1570–1577. doi:10.1097/ACM.0b013e3182a34b05

29. Monrouxe L V., Bullock A, Gormley G, et al. New graduate doctors’ preparedness for practice: A multistakeholder, multicentre narrative study. BMJ Open. 2018;8(8). doi:10.1136/bmjopen-2018-023146

30. Morrow G, Johnson N, Burford B, et al. Preparedness for practice: The perceptions of medical graduates and clinical teams. Med Teach. 2012;34(2):123–135. doi:10.3109/0142159X.2012.643260

31. Kellett J, Papageorgiou A, Cavenagh P, Salter C, Miles S, Leinster SJ. The preparedness of newly qualified doctors - Views of Foundation doctors and supervisors. Med Teach. 2015;37(10):949–954. doi:10.3109/0142159X.2014.970619

32. Cruess RL, Cruess SR, Boudreau JD, Snell L, Steinert Y. Reframing Medical Education to Support Professional Identity Formation. Acad Med. 2014;89(11):1446–1451. doi:10.1097/ACM.0000000000000427

33. Brown MEL, Laughey W, Tiffin PA, Finn GM. Forging a new identity: a qualitative study exploring the experiences of UK-based physician associate students. BMJ Open. 2020;10:33450. doi:10.1136/bmjopen-2019-033450

34. Li Z, Zuo Q, Cheng J, et al. Coronavirus disease 2019 pandemic promotes the sense of professional identity among nurses. Nurs Outlook. October 2020. doi:10.1016/J.OUTLOOK.2020.09.006

35. Wald HS, Anthony D, Hutchinson TA, Liben S, Smilovitch M, Donato AA. Professional Identity Formation in Medical Education for Humanistic, Resilient Physicians. Acad Med. 2015;90(6):753–760. doi:10.1097/ACM.0000000000000725

36. The state of medical education and practice in the UK - GMC.

37. Lefever S, Dal M, Matthíasdóttir Á. Online data collection in academic research: advantages and limitations. Br J Educ Technol. 2007;38(4):574–582. doi:10.1111/j.1467-8535.2006.00638.x

