## Supplemental Survey Questions for "‘I don’t feel like I’m learning how to be a doctor’: early insights regarding the impact of Covid-19 on UK medical student professional identity"

### COVID READY Study: SURVEY OF Volunteering and medical education DURING THE CORONAVIRUS PANDEMIC For All Medical Students in the United Kingdom

#### QUESTIONNAIRE

#### Demographics

- Age – NUMERICAL, PREFER NOT TO SAY
- Gender – MALE, FEMALE, OTHER, PREFER NOT TO SAY
- Medical school – DROP DOWN OF ALL UK MEDICAL SCHOOLS, PREFER NOT TO SAY
- Year at medical school – DROP DOWN 1-6 AND INTERCALATION, PREFER NOT TO SAY
- If you are a final year have you already graduated? YES/NO

#### Previous clinical experience and skills

Have you been employed in a clinical role before?

- Dentist
- Healthcare assistant
- Laboratory scientist
- Paramedic
- Physician’s assistant
- Nurse
- Pharmacist
- Other
- None

What training have you had before? CHECKBOX

- First aid training
- BLS training
- Basic infection control training (e.g. handwashing)
- Surgical scrubbing and gowning
- Infection control training with personal protective equipment (e.g. respirator masks, donning and doffing of goggles and garments)
- Pandemic influenza teaching at medical school
- Disaster medicine teaching at medical school

I am confident independently performing: LIKERT

- Venepuncture
- Cannulation
- Arterial blood gas
- Prescription of medications
- Clerk new admissions (history, examination)
- Organise appropriate investigations for newly clerked patients
- Initiate management plans for newly clerked patients

#### Volunteer to work

Volunteering to work means the recruitment of medical students to work at hospitals in any form during the coronavirus pandemic. Please rate your agreement with the statements below.

Are you aware that medical students may be asked to volunteer to work at hospitals due to the coronavirus pandemic? YES/NO

I have already started as a volunteer in a hospital during the coronavirus pandemic. YES/NO

If you have already volunteered in a hospital, when did you start? DATE

If you have already volunteered, in what capacity have you volunteered?

- Administrative role
- Dentist
- Doctor (interim foundation or equivalent)
- Healthcare assistant
- Laboratory scientist
- Nurse
- Paramedic
- Pharmacist
- Physician’s assistant
- Other

I would be willing to volunteer to work. LIKERT

I believe I will have a positive impact by volunteering to work. LIKERT

Volunteering to work will benefit my medical education. LIKERT

Volunteering to work will benefit my career. LIKERT

Not volunteering will negatively affect my career. LIKERT

I believe medical students should be encouraged to volunteer to work. LIKERT

I believe there are ethical problems with asking medical student to volunteer to work. LIKERT

If you have ethical concerns what are they? FREE TEXT

The coronavirus pandemic has made me consider a career outside of medicine. LIKERT

#### Medical education

I have received sufficient medical education to volunteer to work? LIKERT

Medical school training helped me to understand the strategies taken by Public Health England when controlling the spread of coronavirus. LIKERT

Medical school training has prepared me for conversations about end of life care for patients. LIKERT

My medical education has been negatively affected by the coronavirus. LIKERT

What are the most useful things you learnt at medical school that have prepared you for volunteering? FREE TEXT

Medical school training should include pandemic influenza teaching. LIKERT

I would want additional training if I volunteer to work? LIKERT

#### Motivation/barriers

Why would you volunteer to work? CHECKBOX

- Altruism (e.g. helping those in need)
- Career (e.g. opportunity to improve CV, make new contacts)
- Guilt you would feel if not volunteering
- Moral obligation (e.g. need to do the ‘right’ thing)
- Medical school expectation or directive
- Peer pressure
- Pay
- Professional development and training (e.g. opportunity to learn new skills and gain experience)
- Societal expectations
- Other

Why wouldn’t you volunteer to work? CHECKBOX

- Academic commitments
- Family/social commitments (e.g. caring for a family member)
- Financial implications
- Lack of information on volunteering opportunities available
- Lack of personal protective equipment for healthcare staff
- Personal safety (fear of catching coronavirus)
- Pre-existing health conditions
- Psychological impact
- Work commitments
- Other

#### Role

What role are you willing to do, if you volunteer to work? LIKERT

- Full clinical role expected of a doctor (e.g. clerking, prescribing, ordering investigations, cannulation)
- Assistant medical role (e.g. phlebotomy, cannulation, vaccination)
- Indirect medical care (e.g. providing meals, moving patients)
- Laboratory role (e.g. performing PCR tests)
- Administrative role
- No role

I am willing to do **day** on-calls, if I volunteer to work. LIKERT

I am willing to do **night** on-calls, if I volunteer to work. LIKERT

Would you be willing to do the same role on a ward with coronavirus patients? Yes/No

#### Role for patients with coronavirus (If NO these questions appear)

What role are you willing to do on a ward with coronavirus patients, if you volunteer to work? LIKERT

- Full clinical role expected of a doctor (e.g. clerking, prescribing, ordering investigations, cannulation)
- Assistant medical role (e.g. phlebotomy, cannulation, vaccination)
- Indirect medical care (e.g. providing meals, moving patients)
- Administrative role
- No role

I am willing to do **day** on-calls on a ward with coronavirus patients, if I volunteer to work. LIKERT

I am willing to do **night** on-calls on a ward with coronavirus patients, if I volunteer to work. LIKERT

**Risk**

I am worried about catching coronavirus, if I volunteer to work. LIKERT

If volunteering to work what is your chance of **catching coronavirus** as a percentage? RAW percentage

I feel confident saying no to tasks I am not adequately prepared for. LIKERT

I will receive adequate personal protective equipment if I volunteer to work. LIKERT

I am confident donning and doffing personal protective equipment. LIKERT

I believe I will receive adequate supervision if I volunteer to work. LIKERT

#### Safety

I am aware I should be paid appropriately for any role I undertake in a clinical environment. LIKERT

I am aware I should be provided with a contract prior to starting work. LIKERT

Do you know how to exception report? Yes/No

Do you know how to report clinical incidents e.g. DATIX? Yes/No

Do you know who you should speak to if you have personal, health, or psychiatric problems while volunteering to work? Yes/No

#### Free text questions

What issues are you currently facing as a medical student during the coronavirus pandemic? FREE TEXT

What issues do you think you will encounter if you volunteer to work during the coronavirus pandemic? FREE TEXT

What is the main emotion you feel about working during the coronavirus pandemic? FREE TEXT

Have you heard any stories about medical students volunteering experiences? Please describe them FREE TEXT

Would you like to be followed up for a future questionnaire? This study is part of a longitudinal study evaluating the effect of volunteering. All emails will be anonymised prior to data analysis. EMAIL
